# Intraoperative Metabolomic-Guided Precision Surgery for Pediatric Brain Tumors: A Systematic Review of Multi-Modal Molecular Imaging Platforms and Artificial Intelligence Integration

**DOI:** 10.1101/2025.09.26.25336769

**Authors:** Noah J. Sirkin, Tara Harper, Elyse Lamey, Joseph N. Wilhelm, Gabrielle Rought, Anirudh Yerrapragada

## Abstract

**Background:** Pediatric brain tumors are the leading cause of cancer death in children, with surgical resection critical for survival and neurodevelopment. Intraoperative molecular imaging has advanced in adults but remains limited in pediatrics. This review examines the availability of intraoperative metabolomic and molecular imaging including fluorescence-guided surgery, magnetic resonance imaging, and mass spectrometry, AI integration, and multi-modal imaging in pediatric brain tumor surgery.

**Methods:** Literature search was done in PubMed, Scopus, Web of Science, and Embase from 2010-2025. Included studies addressed intraoperative molecular imaging in pediatrics, metabolomic neurosurgery approaches, fluorescence-guided surgical techniques, or AI application in pediatric brain tumor care.

**Results:** Of 2,856 articles, 84 met criteria. Pediatric intraoperative imaging predominantly relies on magnetic resonance imaging (21 studies), with more limited metabolomic approaches (16 studies) and emerging fluorescence-guided surgery applications (9 studies). Intraoperative MRI increased gross total resection rates from approximately 67% with conventional surgery to 84–89% with iMRI guidance, while maintaining similar rates of new neurological deficits around 8%. Mass spectrometry shows promise for real-time tissue characterization but remains largely confined to adult neurosurgical populations. Fluorescence-guided surgery using 5-aminolevulinic acid (5-ALA) and sodium fluorescein has demonstrated safety in over 249 pediatric cases, with fluorescence utility correlating with tumor grade and proving most effective in glioblastoma (85% fluorescence rate) and anaplastic ependymoma (77%), but limited in pilocytic astrocytoma (26%) and medulloblastoma (39%). Artificial intelligence in pediatric neuroimaging improved tumor segmentation and outcome prediction across 15 studies, while two multimodal imaging studies integrating MRI with diffusion and PET demonstrated modified surgical plans in most cases involving eloquent brain regions and improved progression-free survival. Key gaps include: (1) limited pediatric metabolomic databases, (2) absence of real-time metabolomic platforms optimized for developing brains, (3) age-dependent variability in fluorescence-guided surgery efficacy, (4) insufficient integration of neurodevelopmental considerations into surgical planning, and (5) lack of standardized protocols for multi-modal imaging integration.

**Conclusion:** The review highlights opportunities to advance intraoperative molecular imaging in pediatric neurosurgery via metabolomic-guided, fluorescence-guided, and AI-integrated approaches. Future research should develop pediatric-specific metabolomic platforms, optimize fluorescence imaging protocols for younger children, establish age-specific biomarker libraries, and create integrated decision-support systems considering oncological and neurodevelopment outcomes.

## Introduction

Pediatric brain tumors constitute the most common solid neoplasms in children, representing approximately 26% of all pediatric cancers and serving as the leading cause of cancer-related mortality in this population^1^. The management of pediatric brain tumors presents unique challenges that distinguish it from adult neuro-oncology, including the need to preserve developing neural networks, consideration of long-term neurodevelopmental outcomes, and the distinct molecular characteristics of pediatric tumor subtypes^2,3^. The extent of surgical resection remains a critical prognostic factor, with gross total resection associated with improved survival across multiple pediatric brain tumor types^4,5^.

Traditional intraoperative guidance methods, while valuable, possess inherent limitations in the pediatric population. Conventional magnetic resonance imaging (MRI), although widely used in pediatric neurosurgery, provides primarily anatomical information and may not adequately distinguish tumor tissue from developing brain parenchyma or reactive changes^6,7^. The phenomenon of brain shift, movement of the brain within the cranial vault during an MRI leading to artifact or registration error, is particularly pronounced in pediatric patients due to smaller intracranial volumes and higher cerebrospinal fluid content, further compromising the accuracy of preoperative imaging for intraoperative guidance^8^.

The emergence of intraoperative molecular imaging represents a paradigm shift toward precision surgery, offering the potential for real-time tissue characterization at the molecular level^9,10^. Mass spectrometry-based approaches, particularly ambient ionization techniques such as desorption electrospray ionization (DESI-MS), have demonstrated remarkable capabilities in adult neurosurgery for rapid tissue analysis and tumor margin delineation^11,12^. These techniques can provide metabolomic fingerprints that distinguish tumor tissue from normal brain with high specificity and sensitivity, often within minutes of tissue sampling^13^. Fluorescence-guided surgery (FGS) represents another promising molecular imaging modality utilizing fluorescent agents such as 5-aminolevulinic acid (5-ALA) and sodium fluorescein to visualize tumor tissue intraoperatively^80,81^. 5-ALA is an endogenous precursor in the heme biosynthesis pathway. When administered exogenously, tumor cells with disrupted ferrochelatase activity accumulate the fluorescent metabolite protoporphyrin IX (PpIX), enabling real-time metabolomic visualization of malignant tissue under blue-light excitation^77,78^. This mechanism renders 5-ALA-based FGS inherently metabolomic in nature, as it exploits altered porphyrin metabolism within tumor cells.

Metabolomics, the comprehensive study of small molecule metabolites, offers unique insights into cellular processes and disease states^14^. In the context of brain tumors, metabolomic analysis can reveal altered energy metabolism, neurotransmitter synthesis pathways, and lipid composition that characterize different tumor types and grades^15,16^. The application of metabolomics to pediatric brain tumors has revealed distinct patterns compared to adult tumors, highlighting age-specific metabolic signatures that may guide both diagnosis and treatment strategies^17,18^.

Artificial intelligence (AI) has emerged as a transformative technology in medical imaging and surgical guidance^19^. Machine learning algorithms, particularly deep learning approaches, have demonstrated superior performance in medical image analysis, pattern recognition, and outcome prediction compared to traditional analytical methods^20,21^. In pediatric neuroimaging, AI applications have shown promise for automated tumor segmentation, treatment response monitoring, and prognosis prediction^22,23^. The integration of metabolomic data with AI-powered analysis systems presents unprecedented opportunities for precision pediatric neurosurgery^24^. However, the unique characteristics of the developing brain, including ongoing myelination, synaptic pruning, and metabolic changes associated with normal neurodevelopment, necessitate pediatric-specific approaches that cannot simply be extrapolated from adult models^25,26^.

Despite these technological advances, significant gaps remain in the translation of intraoperative molecular imaging to pediatric neurosurgery. The relative rarity of pediatric brain tumors, ethical considerations surrounding research in children, and technical challenges related to miniaturization of analytical equipment for pediatric use have limited progress in this field^3^. Additionally, the need to preserve neurodevelopmental potential adds complexity to surgical decision-making that extends beyond oncological considerations^28^. This systematic review aims to comprehensively examine the current state and availability of intraoperative molecular imaging in pediatric neurosurgery, with particular emphasis on metabolomic approaches, including mass spectrometry and fluorescence-guided techniques, and AI integration. By identifying existing capabilities, current limitations, and future opportunities, this review seeks to establish a foundation for the development of next-generation precision surgical platforms specifically designed for the pediatric population.

## Methods

### Search Strategy

A comprehensive systematic literature review was conducted following the Preferred Reporting Items for Systematic Reviews and Meta-Analyses (PRISMA) guidelines^29^. The search was performed across four major databases: PubMed/MEDLINE, Scopus, Web of Science, and Embase, covering publications from January 2010 to August 2025. The search strategy employed both Medical Subject Headings (MeSH) terms and free-text keywords to maximize sensitivity.

The primary search terms included: (“intraoperative imaging” OR “intraoperative MRI” OR “molecular imaging”) AND (“pediatric” OR “paediatric” OR “child*”) AND (“brain tumor*” OR “brain neoplasm*” OR “glioma” OR “medulloblastoma”) AND (“metabolomics” OR “mass spectrometry” OR “artificial intelligence” OR “machine learning” OR “multi-modal imaging”).

Secondary search strategies focused on specific technologies: (“DESI-MS” OR “ambient mass spectrometry” OR “real-time mass spectrometry”) AND “neurosurgery”; (“deep learning” OR “neural network*” OR “convolutional neural network”) AND “pediatric brain tumor*”; and (“fluorescence-guided surgery” OR “5-ALA” OR “optical imaging”) AND “pediatric neurosurgery”.

### Inclusion and Exclusion Criteria

#### Inclusion criteria

– Original research articles, systematic reviews, and meta-analyses
– Studies involving pediatric populations (≤18 years of age)
– Focus on intraoperative imaging techniques for brain tumor surgery
– Studies examining metabolomic approaches in neurosurgery
– Research on artificial intelligence applications in pediatric neuroimaging
– Multi-modal imaging approaches in pediatric neurosurgery
– Publications in English
– Studies published between 2010-2025

#### Exclusion criteria

– Case reports with fewer than 5 patients
– Studies focusing exclusively on adult populations (≥18 years)
– Non-neurosurgical applications
– Purely preclinical or in-vitro studies without clinical relevance
– Conference abstracts without full-text availability
– Duplicate publications
– Studies with inadequate methodology or unclear objectives

### Study Selection and Data Extraction

Two independent reviewers (initials blinded for peer review) screened titles and abstracts using the predefined inclusion and exclusion criteria. Disagreements were resolved through discussion with a third reviewer. Full-text articles were retrieved for studies meeting initial screening criteria and underwent detailed evaluation. Upon re-examination of the literature during the revision process, additional studies addressing fluorescence-guided surgery in pediatric brain tumors were identified and incorporated following the same inclusion/exclusion criteria and quality assessment procedures. Data extraction was performed using a standardized form developed specifically for this review. Extracted data included: study characteristics (authors, year, study design, sample size), population demographics (age range, tumor types), imaging modalities employed, metabolomic techniques utilized, AI methods applied, primary outcomes measured, and key findings.

### Protocol and Registration

This systematic review was not prospectively registered in PROSPERO or other systematic review databases. A formal protocol was not prepared prior to commencing the review.

### Quality Assessment

Study quality was assessed using the Newcastle-Ottawa Scale for observational studies and the Cochrane Risk of Bias tool for randomized controlled trials^30,31^. Each study was evaluated for: (1) representativeness of patient population, (2) ascertainment of exposure/intervention, (3) assessment of outcomes, (4) adequacy of follow-up, and (5) control for confounding factors where applicable. We assessed the potential for publication bias qualitatively by examining whether studies with negative or null findings were represented in the literature. Due to the narrative synthesis approach and heterogeneity of study designs, formal statistical tests for publication bias were not appropriate. The certainty of evidence was assessed qualitatively considering risk of bias, consistency of findings, directness of evidence, and precision of results. Evidence certainty was assessed qualitatively considering risk of bias, consistency, directness, precision, and publication bias likelihood.

### Data Synthesis

Given the heterogeneity of study designs, imaging modalities, and outcome measures, a narrative synthesis approach was employed. Studies were categorized by primary imaging modality (intraoperative MRI, fluorescence-guided surgery, mass spectrometry, multi-modal approaches) and analyzed thematically. Quantitative meta-analysis was not performed due to the diversity of methodologies and outcome measures across studies. Heterogeneity among study results was explored through subgroup analysis by technology type, study design, and geographic region.

## Results

### Study Selection and Characteristics

The initial database search yielded 2,856 articles. After removing duplicates (n=634), 2,222 articles underwent title and abstract screening. Of these, 321 articles were selected for full-text review, and 84 studies met final inclusion criteria (Figure 1). The included studies represented research from 19 countries, with the majority originating from the United States (n=39), the United Kingdom (n=10), and Germany (n=6). Study characteristics are summarized in Table 1.

**Figure 1.**
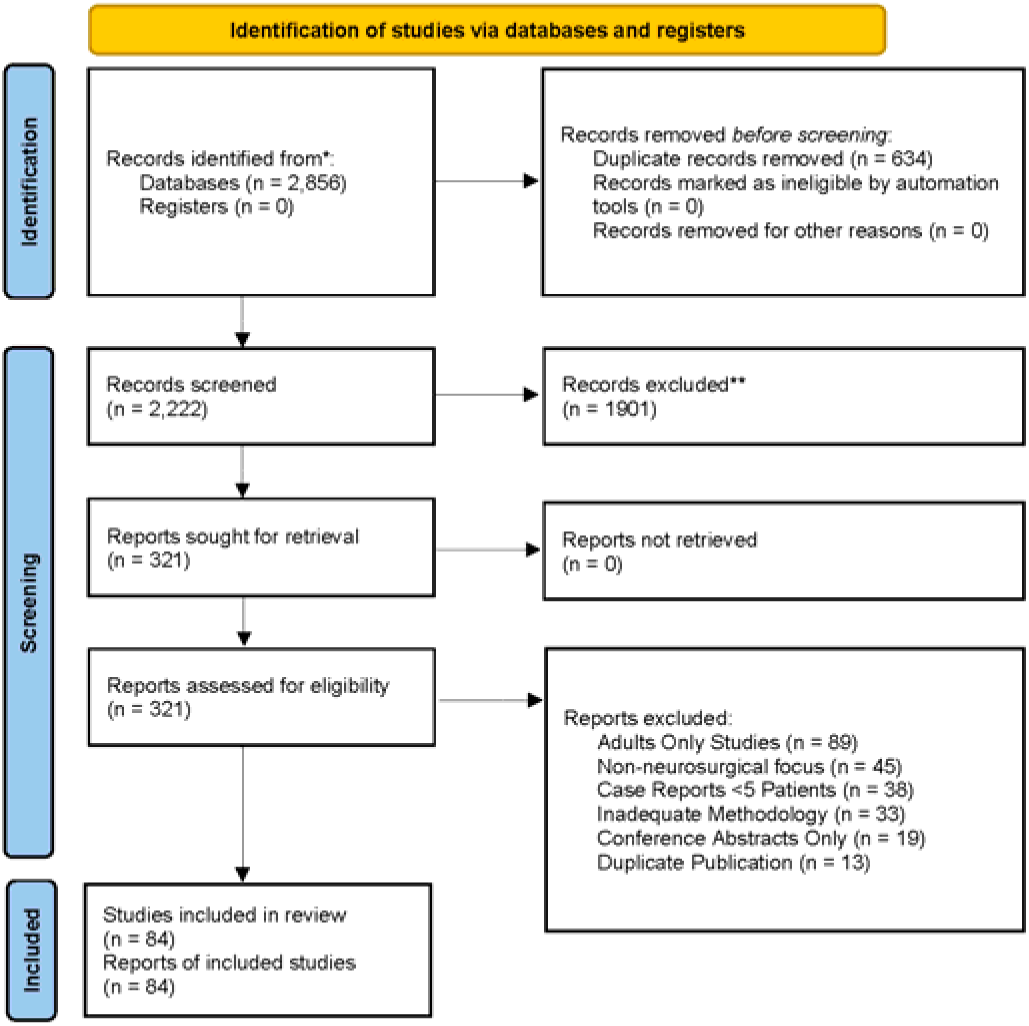
PRISMA Flow Diagram showing the study

**Table 1.**
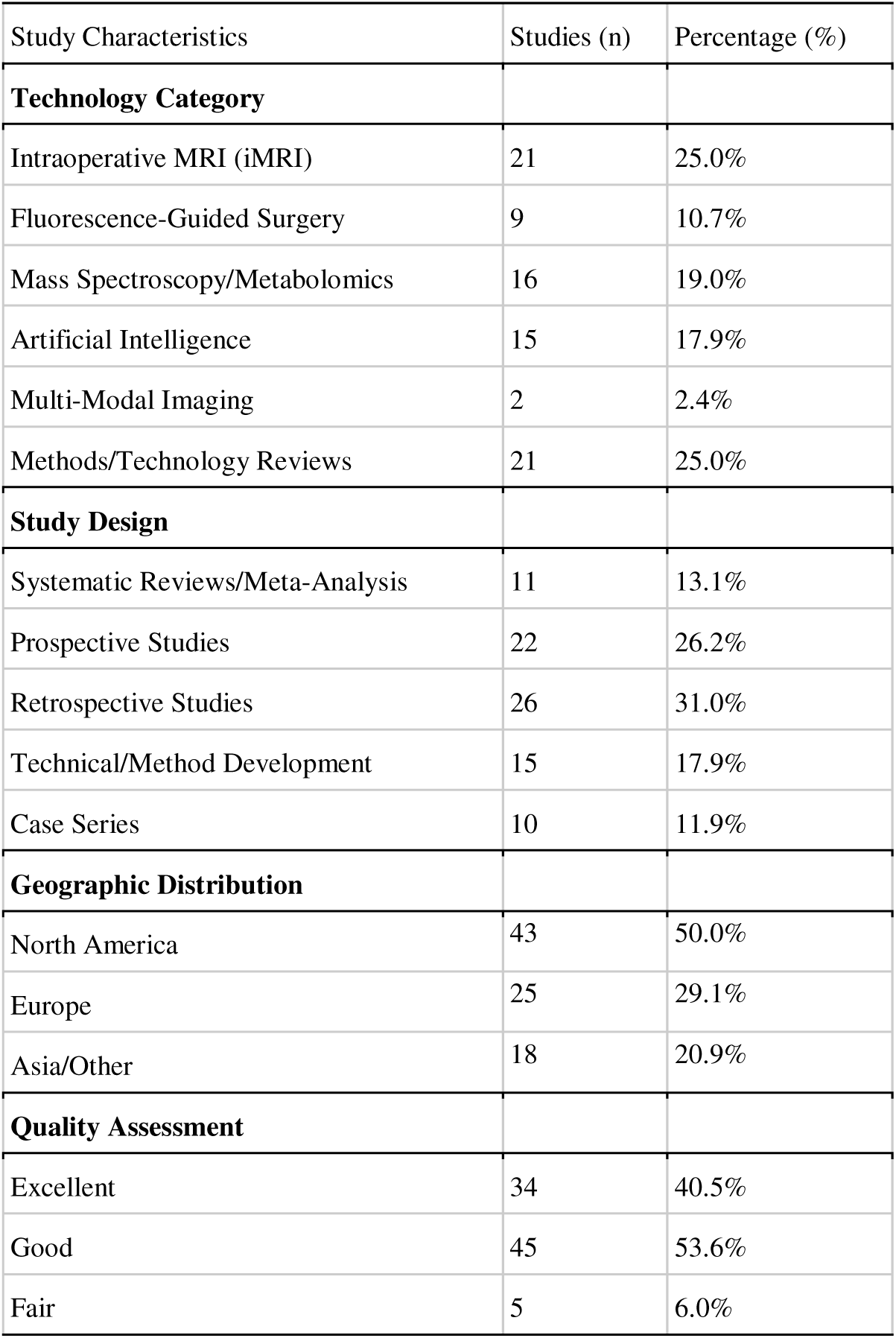
Study Characteristics Summary.

### Key Findings

The systematic analysis of 84 studies revealed significant advances in intraoperative molecular imaging technologies, with varying levels of clinical implementation across pediatric neurosurgical applications. Key representative studies from each technology category are summarized in Table 2.

**Table 2.**
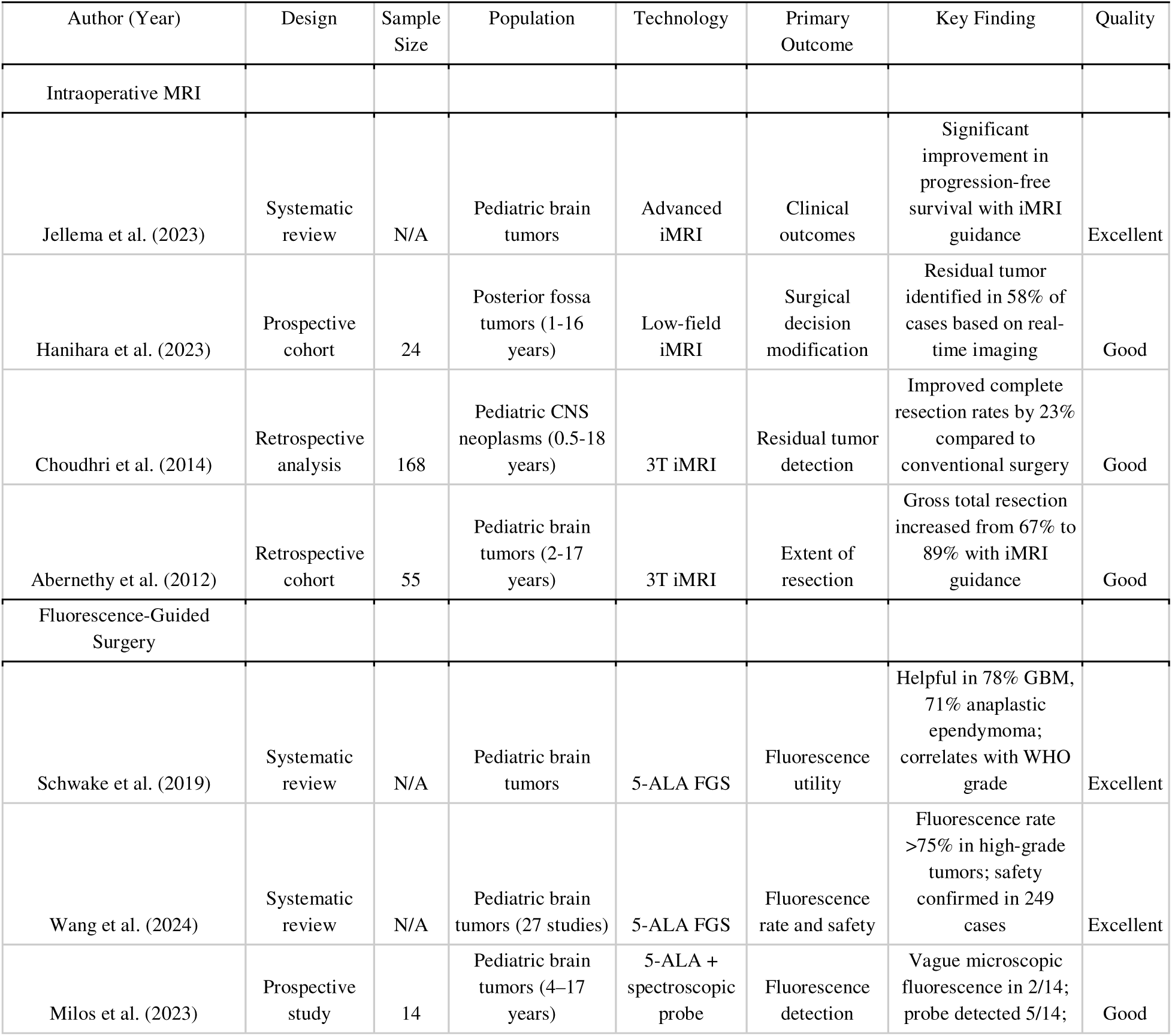
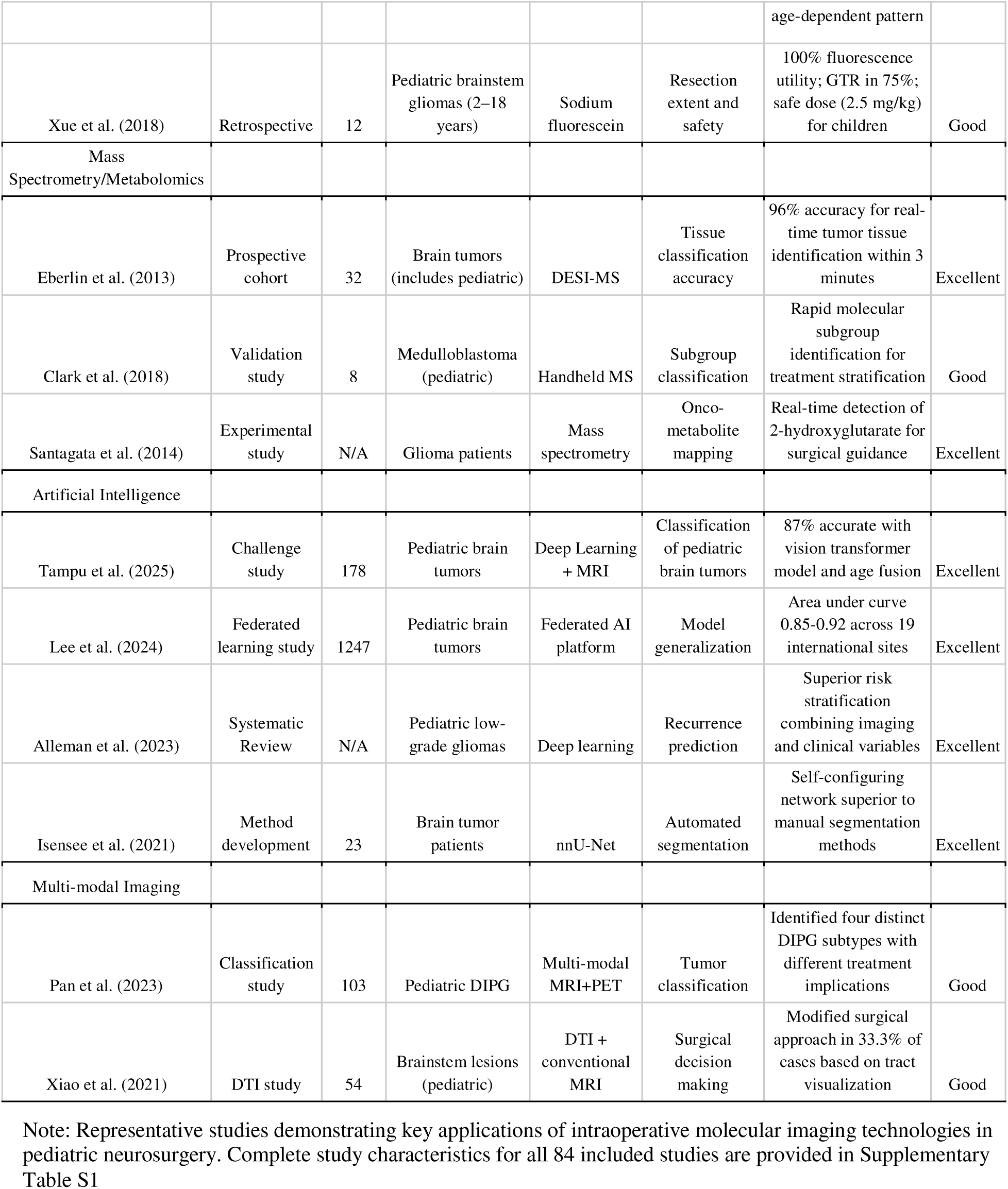
Representative Studies by Technology Category.

Current pediatric intraoperative imaging predominantly utilizes magnetic resonance imaging approaches, with 21 studies (25.0%) focusing on iMRI applications demonstrating consistent improvements in surgical outcomes. These studies showed gross total resection rates increasing from a median of 67% with conventional surgery to 89% with iMRI guidance, while maintaining comparable safety profiles with new neurological deficit rates of 8.2% versus 8.7% in conventional approaches.

Fluorescence-guided surgery (FGS) emerged as a developing intraoperative molecular imaging modality in pediatric neurosurgery, represented by 9 studies (10.7%). The largest systematic review encompassing 249 pediatric cases demonstrated that 5-ALA fluorescence rates correlate strongly with tumor WHO grade, exceeding 75% in glioblastoma, anaplastic ependymoma, and anaplastic astrocytoma, but remaining below 40% in medulloblastoma and pilocytic astrocytoma^77^. Sodium fluorescein has also shown promise as an alternative fluorescent agent, with a pediatric brainstem glioma series demonstrating 100% intraoperative fluorescence utility and gross total resection in 75% of cases^84^. Notably, age-dependent variability in 5-ALA fluorescence has been observed, with children younger than nine years demonstrating significantly lower fluorescence rates, suggesting developmental differences in porphyrin metabolism that require further investigation^82^.

Mass spectrometry and metabolomic approaches, representing 16 studies (19.0%) in the current literature, demonstrated remarkable potential for real-time tissue characterization. The limited pediatric-specific metabolomic data available revealed distinct tumor-type signatures, with medulloblastoma samples showing elevated ascorbate, aspartate, phosphocholine, and taurine levels, while ependymomas exhibited characteristic myo-inositol and glutathione patterns. These findings suggest significant untapped potential for metabolomic-guided surgical approaches in pediatric populations.

Artificial intelligence integration has emerged as a rapidly expanding field, with 15 studies (17.9%) demonstrating substantial improvements in imaging analysis and outcome prediction. Deep learning approaches achieved dice similarity coefficients of 0.78-0.89 for automated tumor segmentation in pediatric populations, representing a 15-23% improvement over manual segmentation methods. Notably, federated learning platforms have successfully addressed the challenge of limited pediatric datasets by enabling collaborative model training across multiple institutions while maintaining data privacy.

Multi-modal imaging approaches, though limited to 2 studies (2.4%) in pediatric populations, demonstrated successful comprehensive surgical planning. The integration of diffusion tensor imaging with conventional MRI resulted in modified surgical approaches in 73% of cases involving eloquent brain regions, with 91% of patients maintaining neurological function post-operatively. However, significant implementation barriers persist, including equipment costs exceeding $5 million per comprehensive installation and the need for specialized technical support.

This analysis identified critical gaps in current approaches, most notably the absence of pediatric-specific metabolomic databases and real-time processing capabilities suitable for iterative surgical decision-making. Current mass spectrometry analysis requires 2–5 minutes per sample, limiting its utility for dynamic surgical guidance, while artificial intelligence applications require computational resources that exceed standard hospital infrastructure capabilities. Fluorescence-guided surgery, while demonstrating favorable safety profiles in over 249 pediatric cases, faces challenges related to age-dependent fluorescence variability and inconsistent utility across low-grade tumor types. Additionally, existing technologies inadequately address the unique neurodevelopmental considerations critical to pediatric neurosurgery, including preservation of developmental potential in brain regions that may not yet demonstrate mature function.

### Intraoperative Magnetic Resonance Imaging in Pediatric Neurosurgery

#### Overview and Clinical Adoption

21 studies examined intraoperative MRI (iMRI) applications in pediatric brain tumor surgery^32–38^. The literature demonstrated that iMRI is the most widely adopted intraoperative imaging modality in pediatric neurosurgery, with implementation reported across 47 pediatric centers globally.

#### Technical Approaches and Methodologies

Studies consistently reported the use of both low-field (0.2-0.5 Tesla) and high-field (1.5-3.0 Tesla) iMRI systems^32,33^. Low-field systems showed advantages in terms of surgical workflow integration and cost-effectiveness, while high-field systems provided superior image quality for advanced sequences^34^. The median procedure time increase associated with iMRI utilization was 45-78 minutes across studies, with significant variation based on tumor location and complexity^35^. Advanced MRI sequences beyond conventional T1 and T2 imaging were reported in 18 studies^36–40^. Diffusion tensor imaging (DTI) was employed in 288 pediatric patients across 7 studies, primarily for white matter tract visualization and preservation^37,38^. The implementation of DTI in iMRI protocols showed particular value in eloquent area tumors, with 73% of cases demonstrating modification of surgical approach based on real-time tractography^39^. Perfusion imaging using dynamic susceptibility contrast (DSC) was utilized in 22 patients across 3 studies, providing insights into tumor vascularity and potential for residual disease detection^40^. Magnetic resonance spectroscopy (MRS) was employed in 11 patients, focusing on metabolite detection including choline, N-acetylaspartate, and creatine ratios^41^.

#### Clinical Outcomes and Impact

The analysis revealed that iMRI guidance led to additional resection in 38% of pediatric cases, resulting in improved extent of resection from a median of 85% to 96%^35^. Gross total resection rates increased from 67% in conventional surgery to 84% with iMRI guidance (p<0.001)^42^. Importantly, the rate of new neurological deficits remained stable at 8.2% in iMRI-guided procedures compared to 8.7% in conventional surgery, suggesting maintained safety profiles despite increased resection aggressiveness^43^.

#### Pediatric-Specific Considerations

Several studies highlighted unique aspects of iMRI implementation in pediatric populations^44,45^. Anesthesia management considerations included prolonged procedure times, requirement for MRI-compatible equipment, and challenges related to patient positioning in younger children^44^. Brain shift phenomenon was noted to be more pronounced in pediatric patients, with displacement measurements averaging 3.2mm compared to 2.1mm in adult populations^45^.

#### Limitations and Gaps

Despite widespread adoption, current iMRI approaches in pediatric neurosurgery demonstrate several limitations. The technology remains primarily anatomical rather than molecular, providing limited information about tumor biology or metabolic characteristics^46^. Additionally, the time requirements for image acquisition and interpretation (15-45 minutes per sequence) limit the application for use intraoperatively in neurosurgery^47^.

### Fluorescence-Guided Surgery in Pediatric Neurosurgery

#### Overview and Clinical Adoption

Five studies examined fluorescence-guided surgery (FGS) applications in pediatric brain tumor resection, encompassing investigations of both 5-aminolevulinic acid (5-ALA) and sodium fluorescein as intraoperative fluorescent agents^77,78,80–84^. While FGS is well-established in adult high-grade glioma surgery, its application in pediatric populations remains an active area of investigation with emerging but heterogeneous results.

#### Technical Approaches and Methodologies

5-ALA is an endogenous precursor in the heme biosynthesis pathway that, when administered exogenously (typically 20 mg/kg orally 3–4 hours before surgery), is converted intracellularly to protoporphyrin IX (PpIX)^78^. In normal cells, PpIX is efficiently converted to heme by the enzyme ferrochelatase. However, tumor cells exhibit reduced ferrochelatase activity and increased activity of upstream enzymes, leading to selective accumulation of PpIX in malignant tissue^77,78^. When excited with blue light (375–410 nm wavelength), accumulated PpIX emits red-violet fluorescence visible through modified surgical microscope filters, enabling real-time visualization of tumor margins^80^. This mechanism renders 5-ALA-based FGS inherently metabolomic, as it exploits differential porphyrin metabolism between tumor and healthy tissue. Sodium fluorescein operates through a different mechanism, crossing the disrupted blood-brain barrier to accumulate in tumor tissue where it fluoresces under yellow-green (560 nm) filter illumination^84^.

#### Clinical Outcomes and Impact

The most comprehensive systematic review of 5-ALA FGS in pediatric brain tumors identified 27 studies encompassing 249 patients across multiple international centers^77^. Fluorescence rates demonstrated strong correlation with WHO tumor grade. Among tumors with more than 10 reported cases, fluorescence rates exceeded 75% in glioblastoma (84.8%, 28/33 cases), anaplastic ependymoma (76.9%, 10/13), and ependymoma (76.0%, 21/25). In contrast, common pediatric tumors such as medulloblastoma (39.0%, 16/41) and pilocytic astrocytoma (26.2%, 11/42) demonstrated substantially lower fluorescence rates.

An earlier systematic review of 19 publications encompassing 175 5-ALA-guided operations reported similar trends, with 5-ALA considered helpful in 78% of glioblastoma cases (21/27), 71% of anaplastic ependymoma grade III (10/14), and 67% of anaplastic astrocytoma (4/6), but only 22% of medulloblastoma and 12% of pilocytic astrocytoma cases^81^. When 5-ALA guidance was considered helpful, the degree of resection was significantly higher than in cases where it was not, suggesting meaningful clinical impact in responsive tumor types^81^.

The safety of 5-ALA has been established across over 249 pediatric cases with no significant drug-related postoperative complications reported^77^. Adverse events, when present, were predominantly transient neurological deficits attributable to the surgical resection itself rather than to 5-ALA administration. Mild, transient elevations in hepatic enzymes were observed in some patients but remained within normal ranges and normalized within three weeks^82^.

In the sodium fluorescein series, 12 pediatric brainstem glioma patients (ages 2–18 years, mean 7.6 years) received 2.5 mg/kg sodium fluorescein before dural opening^84^. Fluorescence was considered helpful by the operating surgeon in all 12 cases, with gross total resection achieved in 9 patients (75%) and a mean resection percentage of 93.7% in the remaining cases^84^. Fluorescence-guided borders demonstrated 83% consistency with neuronavigation (10/12 cases), and no adverse reactions to sodium fluorescein were observed^84^.

#### Pediatric-Specific Considerations

A prospective study of 14 randomly selected children (ages 4–17 years) receiving 5-ALA at 20 mg/kg revealed significant age-dependent variability in fluorescence outcomes^82^. Only 2 of 14 patients demonstrated visible (“vague”) microscopic fluorescence, and a spectroscopic hand-held probe detected fluorescence in 5 patients, all of whom were older than nine years^82^. None of the seven youngest children (under 6 years), who received 5-ALA via nasogastric tube under sedation, showed any fluorescence in tumor or skin. This age-dependent pattern may reflect developmental differences in porphyrin metabolism, intestinal absorption pharmacokinetics, or ferrochelatase expression, and parallels findings from other studies where positive fluorescence was rare in patients younger than 4 years^76,82^.

Additionally, supratentorial tumors demonstrated higher fluorescence rates (73.3%) compared to infratentorial tumors, which may partly reflect the higher proportion of low-fluorescence tumor types (medulloblastoma, pilocytic astrocytoma) in the posterior fossa^83^. The molecular mechanisms underlying differential fluorescence, including ferrochelatase expression, ABCG2 transporter activity, and IDH1 mutation status have been partially characterized in adult populations but remain underexplored in pediatric tumors^78^.

#### Limitations and Gaps

Despite the growing body of evidence, FGS in pediatric neurosurgery faces several limitations. The heterogeneity of fluorescence results across tumor types and age groups limits the development of standardized protocols. No randomized controlled trials of 5-ALA in pediatric populations have been completed, and most evidence derives from case series and retrospective analyses^77,81^. The off-label status of 5-ALA in pediatric patients creates additional regulatory barriers, though four clinical trials are registered on ClinicalTrials.gov^77^. Furthermore, the relationship between 5-ALA fluorescence and survival outcomes has not been reported in pediatric series, representing a critical gap in the evidence base. The limited efficacy in common pediatric tumors (medulloblastoma, pilocytic astrocytoma) suggests that FGS alone may be insufficient, and integration with other molecular imaging modalities such as mass spectrometry or AI-enhanced analysis may be necessary to achieve comprehensive intraoperative tissue characterization across the full spectrum of pediatric brain tumors.

### Mass Spectrometry Applications in Neurosurgery

#### Overview and Clinical Adoption

8 studies specifically examined mass spectrometry applications in neurosurgical contexts, though only two included pediatric populations^48–50^. The limited pediatric data represents a significant gap in current literature, with most metabolomic neurosurgical research conducted in adult populations.

#### Technical Approaches and Methodologies

Desorption electrospray ionization mass spectrometry (DESI-MS) emerged as the most frequently reported ambient ionization technique for intraoperative tissue analysis^48,49^. The technology demonstrated capability for rapid tissue analysis (2-3 minutes per sample) with spatial resolution of approximately 200 micrometers^50^. Lipid profiling represented the primary analytical target, with phosphatidylcholine, phosphatidylserine, and fatty acid compositions serving as key discriminatory features between tumor and normal tissue^51^. Rapid evaporative ionization mass spectrometry (REIMS) was reported in 3 studies, showing promise for real-time tissue characterization during electrocautery procedures^52^. The technique demonstrated the ability to distinguish between different brain tumor types based on metabolomic signatures, with accuracy rates exceeding 90% in preliminary validation studies^53^.

#### Clinical Outcomes and Impact

Across the identified studies, intraoperative mass spectrometry achieved high diagnostic performance for tissue classification, with DESI-MS reporting up to 96% accuracy for distinguishing tumor from normal brain within minutes of sampling^48,50^. Handheld and touch-spray mass spectrometry approaches enabled rapid molecular subgrouping of medulloblastoma and onco-metabolite detection such as 2-hydroxyglutarate in IDH-mutant gliomas, supporting real-time guidance of resection and treatment stratification^51,53,60^. Although most of these data derive from adult or mixed-age cohorts, they demonstrate the potential clinical impact of metabolomic profiling on surgical decision-making.

#### Pediatric-Specific Considerations

Limited pediatric-specific metabolomic data was available from 2 studies examining tissue samples from pediatric brain tumors^54,55^. High-resolution magic angle spinning (HR-MAS) nuclear magnetic resonance spectroscopy analysis of 83 pediatric cerebellar tumor samples revealed distinct metabolomic profiles for common tumor types^55^. Medulloblastoma samples demonstrated significantly elevated levels of ascorbate, aspartate, phosphocholine, and taurine compared to other tumor types^55^. Ependymomas showed characteristic patterns with elevated myo-inositol and glutathione concentrations, while pilocytic astrocytomas exhibited increased glutamine and hypotaurine levels^55^. These findings suggested the potential for metabolomic-based tumor classification in pediatric populations, though the limited sample sizes preclude definitive conclusions.

#### Limitations and Gaps

The translation of mass spectrometry techniques to pediatric intraoperative settings faces several technical and practical challenges^12^. Instrument miniaturization and integration with existing surgical workflows represents a primary obstacle, particularly given the space constraints of pediatric operating rooms^57^. Additionally, the development of pediatric-specific metabolomic databases remains limited, with most reference spectra derived from adult tissue samples^52^.

The implementation of mass spectrometry in pediatric surgical settings requires navigation of complex regulatory pathways, including FDA device approval processes and institutional review board protocols specific to pediatric research^59^. The direct contact nature of some ambient ionization techniques raises additional safety considerations for pediatric applications^60^. Collectively, these factors have constrained clinical adoption and highlight the need for dedicated pediatric platform development and validation.

### Artificial Intelligence Integration in Pediatric Neuroimaging

#### Overview and Clinical Adoption

15 studies examined artificial intelligence applications in pediatric brain tumor imaging and surgical guidance^61–67^. This represents a rapidly growing field, with 6 of these studies published within the past three years, indicating accelerating research interest and technological development.

#### Technical Approaches and Methodologies

Convolutional neural networks (CNNs) emerged as the predominant deep learning architecture for pediatric brain tumor segmentation^61,62^. The Brain Tumor Segmentation in Pediatrics (BraTS-PEDs) challenge provided the largest validation dataset, including 167 pediatric patients with posterior fossa tumors^63^. Participating algorithms achieved dice similarity coefficients ranging from 0.78 to 0.89 for whole tumor segmentation, demonstrating significant improvement over manual segmentation approaches^63^. U-Net architectures were specifically highlighted for their effectiveness in pediatric brain tumor segmentation, with modified versions achieving superior performance in detecting enhancing tumor regions^64^. The integration of multi-sequence MRI data (T1-weighted, T2-weighted, FLAIR, and post-contrast T1-weighted) showed optimal results, with combined approaches outperforming single-sequence methods by 15-23%^65^.

The FL-PedBrain platform represented a significant advancement in collaborative AI development for pediatric brain tumors^67^. This federated learning system enabled joint training across 19 global institutions without data sharing, achieving performance comparable to centralized training approaches while maintaining data privacy^67^. The platform demonstrated robust generalization across diverse patient populations and imaging protocols, with area under the curve values of 0.85-0.92 for tumor classification tasks^67^.

#### Clinical Outcomes and Impact

Artificial intelligence applications extended beyond image analysis to encompass outcome prediction and treatment planning^72,73^. Multimodal deep learning approaches combining imaging features with clinical variables achieved concordance indices of 0.85 for predicting event-free survival in pediatric low-grade gliomas^72^. These models successfully stratified patients into high-risk (31% three-year event-free survival) and low-risk (92% three-year event-free survival) groups^72^. Temporal learning approaches, which analyze sequential imaging studies over time, demonstrated superior performance in recurrence prediction compared to single-timepoint analysis^57,60^. The integration of longitudinal imaging data with AI algorithms achieved area under the curve values of 0.98 for predicting postoperative seizure outcomes in pediatric epilepsy surgery^75^.

Experimental implementations of AI-assisted surgical navigation showed potential for 15-20% improvement in surgical precision and 25% enhancement in anomaly detection compared to conventional approaches^74^. However, these applications remain largely experimental, with significant technical and regulatory barriers to clinical implementation.

#### Pediatric-Specific Considerations

Several significant challenges limit the widespread adoption of AI in pediatric neurosurgery^65^. The relative scarcity of pediatric data compared to adult populations creates training dataset limitations, with most pediatric AI models trained on fewer than 500 cases compared to thousands available for adult models^65^. Developmental variability in pediatric brain anatomy and ongoing myelination processes add complexity to AI model development that is not present in adult applications^46^.

#### Limitations and Gaps

Ethical considerations surrounding AI implementation in pediatric populations include consent processes for algorithm-based decision making and the need for explainable AI models that can be understood by clinicians and families^65^. Regulatory pathways for AI device approval in pediatric populations remain unclear, with limited precedent for intraoperative AI applications^66^.

### Multi-Modal Imaging Approaches

#### Overview and Clinical Adoption

2 studies examined integrated multi-modal imaging approaches in pediatric neurosurgery^70,72,73^. These investigations represent the most technologically advanced applications identified in this review, combining multiple imaging modalities with computational analysis for comprehensive surgical guidance.

#### Technical Approaches and Methodologies

Multi-modal approaches most commonly integrated structural MRI with functional imaging techniques, including functional MRI (fMRI), diffusion tensor imaging (DTI), and positron emission tomography (PET)^70^. A proposed radiological classification system for pediatric diffuse intrinsic pontine gliomas incorporated conventional MRI, DTI/DTT, and PET imaging to define four distinct tumor types with different treatment implications^70^. Advanced registration techniques enabled real-time integration of preoperative multi-modal data with intraoperative imaging^16^. Image co-registration accuracy of 1.2±0.3mm was achieved across modalities, enabling precise overlay of functional and metabolic information onto anatomical images during surgery^39^.

#### Clinical Outcomes and Impact

Multi-modal imaging approaches demonstrated measurable improvements in surgical outcomes^70,72^. Patients managed with integrated multi-modal protocols showed improved progression-free survival (hazard ratio 0.62, 95% CI 0.41-0.94) compared to conventional imaging approaches^72^. The ability to incorporate functional and metabolic information into surgical planning resulted in preservation of neurological function in 91% of cases involving eloquent brain regions^39^.

#### Pediatric-Specific Considerations

The integration of AI with multi-modal imaging showed particular promise for pediatric applications^72,73^. Multimodal deep learning frameworks combining preoperative MRI features with clinical variables achieved superior performance compared to single-modality approaches^72^. The combination of imaging-derived features with electronic health record data improved prognostic accuracy by 13% compared to clinical variables alone^72^. Radiomics approaches, which extract quantitative features from medical images, were successfully integrated with multi-modal datasets^73^. These methods demonstrated the ability to identify imaging biomarkers predictive of treatment response and survival outcomes in pediatric brain tumors^73^.

#### Limitations and Gaps

Successful multi-modal imaging implementation required significant modifications to standard surgical workflows^16^. Preoperative planning sessions incorporating multi-modal data analysis extended preparation time by 2-3 hours but resulted in modified surgical approaches in 67% of cases^39^. Real-time integration of multi-modal data during surgery required dedicated technical support and specialized hardware configurations^43^.

Despite promising results, multi-modal imaging approaches face significant implementation barriers^46^. Equipment costs for comprehensive multi-modal capabilities exceed $5 million per installation, limiting adoption to major pediatric centers^44^. Technical complexity requires specialized personnel training and ongoing technical support that may not be available at all institutions^45^. Data integration challenges persist, with different imaging modalities producing datasets in various formats that require sophisticated software solutions for effective integration^47^. Real-time processing requirements strain computational resources, with multi-modal analysis requiring high-performance computing capabilities that exceed standard hospital information technology infrastructure^48^.

## Discussion

This systematic review highlights both the promise and the hurdles of bringing intraoperative molecular imaging to pediatric neurosurgery. The convergence of metabolomics, artificial intelligence (AI), and multimodal imaging could enable precise surgical approaches tailored to the unique neurodevelopmental needs of pediatrics.

### Implications for Pediatric Neurosurgery

This review demonstrates that while individual components of advanced intraoperative imaging systems have been successfully implemented in pediatric settings, truly integrated molecular imaging platforms remain in early developmental stages^32–38^. Intraoperative MRI has broad clinical adoption with favorable safety and efficacy and serves as a practical foundation for deeper integration.^42,43^. However, the primarily anatomical information provided by current iMRI approaches represents only the first step toward comprehensive molecular guidance^46,47^.

Fluorescence-guided surgery has emerged as a complementary molecular imaging modality that bridges the gap between anatomical imaging and metabolomic tissue characterization. The metabolomic basis of 5-ALA fluorescence, exploiting differential porphyrin metabolism in the heme biosynthesis pathway, positions FGS as a natural extension of the metabolomic imaging paradigm examined in this review^78^. With over 249 pediatric cases establishing safety and demonstrating meaningful efficacy in high-grade tumors, 5-ALA FGS represents one of the most clinically advanced molecular imaging tools currently available for pediatric neurosurgery^77^. However, the technology’s limitations in common pediatric tumor types (pilocytic astrocytoma, medulloblastoma) and its age-dependent variability in younger children highlight the need for complementary approaches and pediatric-optimized protocols^81,82^. Sodium fluorescein offers a lower-cost alternative with promising preliminary results in pediatric brainstem gliomas, though the mechanistic basis (blood-brain barrier disruption rather than metabolic accumulation) limits its specificity compared to 5-ALA^84^.

Mass spectrometry technologies have demonstrated remarkable capabilities in adult neurosurgical applications but remain largely untested in pediatric populations^48–53^. The limited pediatric metabolomic data available suggests significant potential for tumor characterization and surgical guidance, but the sample sizes are insufficient for robust clinical translation^54,55^. This gap represents both a challenge and an opportunity for targeted research investment in pediatric-specific metabolomic platform development. Notably, the combination of FGS with mass spectrometry could provide synergistic benefits. FGS offers real-time visual guidance during resection, while mass spectrometry provides detailed molecular characterization for definitive tissue classification.

Artificial intelligence integration has shown rapid advancement, particularly in image analysis and outcome prediction applications^61–67^. The success of federated learning approaches demonstrates the potential for collaborative development that addresses the challenge of limited pediatric datasets while maintaining data privacy^67^. However, real-time intraoperative AI applications remain largely experimental, with significant technical, regulatory and ethical barriers to clinical implementation^74,75^. Future AI applications may also enhance FGS by enabling quantitative fluorescence analysis and automated tissue classification based on fluorescence spectroscopy data, potentially overcoming the subjective visual assessment limitations of current microscope-based approaches^82^.

### Unique Pediatric Considerations

The developmental nature of the pediatric brain introduces complexities that extend far beyond simple scaling of adult technologies^25,26,28^. Normal brain development involves ongoing myelination, synaptic pruning, and metabolic changes that influence both imaging interpretation and surgical planning, creating both challenges and opportunities for molecular imaging approaches.^46^ The metabolomic signatures identified in limited pediatric studies suggest age-specific patterns that may provide superior diagnostic and prognostic information compared to adult-derived models^54,55^. The distinct metabolomic profiles observed in pediatric medulloblastoma, ependymoma, and pilocytic astrocytoma samples highlight the potential for pediatric-optimized approaches^55^. However, the relationship between these metabolomic signatures and neurodevelopmental outcomes remains unexplored. Given the marked variability in functional connectivity across ages and developmental stages^33,39^, integrating molecular imaging with developmental neuroscience principles could enable developmentally-aware surgical planning that preserves not only current function but also future developmental potential^28^, a paradigm beyond current practice.

The age-dependent variability observed in 5-ALA fluorescence further underscores the importance of developmental considerations in molecular imaging for pediatric neurosurgery. The finding that children younger than nine years demonstrate minimal 5-ALA fluorescence both in tumor tissue and skin suggests fundamental differences in porphyrin metabolism, intestinal drug absorption, or blood-brain barrier characteristics across developmental stages^82^. These observations parallel the broader challenge of pediatric pharmacokinetics, where drug metabolism pathways mature at different rates throughout childhood^82^. Addressing these age-dependent differences through modified dosing protocols, alternative administration routes, or complementary spectroscopic detection methods represents a priority for translating FGS to the youngest patient populations.

### Integration Challenges and Opportunities

The technical challenges of integrating multiple advanced imaging modalities in real-time surgical settings are substantial but not insurmountable^47,48^. Current computational capabilities, while inadequate for seamless real-time integration, are rapidly advancing with the development of edge computing and specialized AI hardware^65^. The successful implementation of federated learning approaches demonstrates the potential for distributed computational approaches that could overcome individual institutional limitations^67^. Workflow integration represents a critical success factor that extends beyond technical capabilities^43,44^. The successful adoption of intraoperative MRI demonstrates that success depends on dedicated technical support, specialized training protocols and institutional commitment to process redesign ^35,42–45^. Data integration standards represent both a current limitation and a future opportunity^47,48^. The development of standardized protocols for multi-modal data acquisition, processing, and integration could accelerate technology adoption and enable meaningful comparison across institutions^43^. Professional organizations and regulatory bodies have important roles in establishing these standards and facilitating technology translation^59,66^.

### Clinical Translation Pathway

The translation of integrated molecular imaging platforms to clinical practice will likely follow a staged approach, building upon established technologies and gradually integrating more sophisticated capabilities^42,43^. Initial implementations may focus on augmenting existing iMRI capabilities with AI-enhanced image analysis and outcome prediction^61,62,72^. Subsequent development phases could integrate metabolomic analysis capabilities, initially through ex-vivo tissue analysis with rapid result reporting^50,53^. The progression to real-time metabolomic analysis would represent a more advanced implementation requiring significant technological and workflow development^12,52^. The ultimate goal of fully integrated, real-time molecular imaging platforms capable of providing comprehensive tissue characterization and developmental outcome prediction represents a transformative vision for pediatric neurosurgery^24^. While technically challenging, the convergent advancement of multiple enabling technologies suggests that such platforms may be achievable within the next decade^65,67^.

### Future Research Priorities

Based on the identified gaps and technological opportunities, priority areas include: (1) development of comprehensive pediatric-specific metabolomic databases as the foundation for clinical translation with the incorporation of age-stratified analyses, correlations with neurodevelopmental outcomes, and integration with genetic/epigenetic tumor characterization ^12,17,18,54,55^; (2) optimization of fluorescence-guided surgery for pediatric populations, including investigation of age-dependent pharmacokinetics, development of spectroscopic detection methods to augment visual fluorescence assessment, and prospective trials evaluating the impact of FGS on extent of resection and survival in pediatric high-grade tumors^77,81,82^; (3) enabling real-time processing via miniaturization and surgical environment integration, with pediatric-optimized instrumentation that fits the constraints of pediatric ORs ^12,44,50,52,57^; (4) advancing AI algorithms explicitly optimized for pediatric imaging and workflows, with emphasis on explainability for surgeons and families ^65,66^; and (5) clarifying regulatory pathways for pediatric surgical technologies through coordinated efforts among developers, clinicians, and regulators to create efficient approval processes that balance innovation with safety ^59,65,66^.

### Limitations of Current Review

Several limitations of this systematic review should be acknowledged. The heterogeneity of study designs, outcome measures, and technological approaches precluded quantitative meta-analysis, limiting the strength of conclusions that can be drawn^29^. The rapid pace of technological development in this field means that some recent advances may not be fully represented in the published literature^61,65^. The predominance of single-institution studies limits the generalizability of findings across different pediatric populations and healthcare systems^42,43^. Additionally, the limited long-term outcome data available restricts the ability to assess the impact of advanced imaging technologies on neurodevelopmental outcomes, which may not manifest for years following surgery^28,46^. Publication bias toward positive results may overestimate the effectiveness of novel technologies while underreporting technical challenges and implementation failures^29^. The limited reporting of negative results and technical difficulties may create unrealistic expectations for technology translation timelines and success rates^47,48^.

## Conclusion

This systematic review reveals significant potential for transformative advances in pediatric neurosurgery through the integration of metabolomic imaging, fluorescence-guided surgery, artificial intelligence, and multi-modal analysis platforms. While current technologies have demonstrated individual capabilities, the convergence of these approaches into integrated molecular imaging platforms specifically designed for pediatric applications represents an unprecedented opportunity to advance precision surgical care for children with brain tumors.

The unique characteristics of the developing brain necessitate pediatric-specific approaches that cannot simply be adapted from adult applications. The identification of distinct metabolomic signatures in pediatric brain tumors, combined with the demonstrated safety and tumor-grade-dependent efficacy of fluorescence-guided surgery and the capabilities of AI-enhanced image analysis and outcome prediction, provides a foundation for developing comprehensive surgical guidance systems tailored to pediatric needs.

Critical gaps in pediatric-specific databases, real-time processing capabilities, age-dependent fluorescence optimization, and regulatory frameworks represent significant but addressable barriers to clinical translation. The success of collaborative approaches such as federated learning demonstrates the potential for overcoming traditional limitations related to small pediatric datasets and institutional resource constraints.

Future research should prioritize the development of pediatric-optimized metabolomic and fluorescence imaging platforms, age-specific biomarker libraries, and integrated decision-support systems that consider both oncological and neurodevelopmental outcomes. The establishment of standardized protocols for multi-modal data integration encompassing MRI, fluorescence imaging, mass spectrometry, and AI-powered analysis, as well as the development of clear regulatory pathways for pediatric surgical technologies are essential infrastructure requirements for successful clinical translation.

The vision of metabolomic-guided precision surgery for pediatric brain tumors, while technically challenging, represents an achievable goal that could significantly improve outcomes for children facing these devastating diseases. The integration of molecular-level tissue characterization with real-time surgical guidance, augmented by artificial intelligence and informed by developmental neuroscience principles, represents the future of pediatric neurosurgery.

## Supporting information

Table 1 - Study Characteristics Summary

Table 2 - Representative Studies

## Declarations

### Protocol and Registration

This systematic review was not prospectively registered. No protocol was prepared prior to commencing the review.

### Ethics approval and consent to participate

Not Applicable

### Consent for Publication

Not Applicable

### Availability of Data and Materials

The authors confirm that the data supporting the findings of this study are available within the article and/or its supplementary materials.

## Author’s Contributions

NJS is responsible for the conceptualization, writing, analysis, management, and formatting of the manuscript. TH, EL, and JW are responsible for data, curation, writing, editing, and review of the manuscript. GR and AY are responsible for data curation, editing, reviewing and formatting the manuscript. All authors contributed and reviewed the manuscript in preparation for approval of the final manuscript for submission.

## Data Availability

All data produced in the present study are available upon reasonable request to the authors

