## Supplementary material for "Intraoperative Metabolomic-Guided Precision Surgery for Pediatric Brain Tumors: A Systematic Review of Multi-Modal Molecular Imaging Platforms and Artificial Intelligence Integration": Table 1 - Study Characteristics Summary

| Study Characteristics | Studies (n) | Percentage (%) |
|---|---|---|
| **Technology Category** | | |
| Intraoperative MRI (iMRI) | 21 | 25.0% |
| Fluoscence-Guided Surgery | 9 | 10.7% |
| Mass Spectroscopy/Metabolomics | 16 | 19.0% |
| Artificial Intelligence | 15 | 17.9% |
| Multi-Modal Imaging | 2 | 2.4% |
| Methods/Technology Reviews | 21 | 25.0% |
| **Study Design** | | |
| Systematic Reviews/Meta-Analysis | 11 | 13.1% |
| Prospective Studies | 22 | 26.2% |
| Retrospective Studies | 26 | 31.0% |
| Technical/Method Development | 15 | 17.9% |
| Case Series | 10 | 11.9% |
| **Geographic Distribution** | | |
| North America | 43 | 50.0% |
| Europe | 25 | 29.1% |
| Asia/Other | 18 | 20.9% |
| **Quality Assessment** | | |
| Excellent | 34 | 40.5% |
| Good | 45 | 53.6% |
| Fair | 5 | 6.0% |
