## Supplementary material for "Intraoperative Metabolomic-Guided Precision Surgery for Pediatric Brain Tumors: A Systematic Review of Multi-Modal Molecular Imaging Platforms and Artificial Intelligence Integration": Table 2 - Representative Studies

**Representative Studies by Technology Category - Table 2**

| Author (Year) | Design | Sample Size | Population | Technology | Primary Outcome | Key Finding | Quality |
|---|---|---|---|---|---|---|---|
| Intraoperative MRI | | | | | | | |
| Jellema et al. (2023) | Systematic review | 847 | Pediatric brain tumors | Advanced iMRI | Clinical outcomes | Significant improvement in progression-free survival with iMRI guidance | Excellent |
| Hanihara et al. (2023) | Prospective cohort | 24 | Posterior fossa tumors (1-16 years) | Low-field iMRI | Surgical decision modification | Residual tumor identified in 58% of cases based on real-time imaging | Good |
| Choudhri et al. (2014) | Retrospective analysis | 168 | Pediatric CNS neoplasms (0.5-18 years) | 3T iMRI | Residual tumor detection | Improved complete resection rates by 23% compared to conventional surgery | Good |
| Abernethy et al. (2012) | Retrospective cohort | 55 | Pediatric brain tumors (2-17 years) | 3T iMRI | Extent of resection | Gross total resection increased from 67% to 89% with iMRI guidance | Good |
| Fluorescence-Guided Surgery | | | | | | | |
| Schwake et al. (2019) | Systematic review | 175 | Pediatric brain tumors | 5-ALA FGS | Fluorescence utility | Helpful in 78% GBM, 71% anaplastic ependymoma; correlates with WHO grade | Excellent |
| Wang et al. (2024) | Systematic review | 249 | Pediatric brain tumors (27 studies) | 5-ALA FGS | Fluorescence rate and safety | Fluorescence rate >75% in high-grade tumors; safety confirmed in 249 cases | Excellent |
| Milos et al. (2023) | Prospective study | 14 | Pediatric brain tumors (4–17 years) | 5-ALA + spectroscopic probe | Fluorescence detection | Vague microscopic fluorescence in 2/14; probe detected 5/14; age-dependent pattern | Good |
| Xue et al. (2018) | Retrospective | 12 | Pediatric brainstem gliomas (2–18 years) | Sodium fluorescein | Resection extent and safety | 100% fluorescence utility; GTR in 75%; safe dose (2.5 mg/kg) for children | Good |
| Mass Spectrometry/Metabolomics | | | | | | | |
| Eberlin et al. (2013) | Prospective cohort | 32 | Brain tumors (includes pediatric) | DESI-MS | Tissue classification accuracy | 96% accuracy for real-time tumor tissue identification within 3 minutes | Excellent |
| Clark et al. (2018) | Validation study | 8 | Medulloblastoma (pediatric) | Handheld MS | Subgroup classification | Rapid molecular subgroup identification for treatment stratification | Good |
| Santagata et al. (2014) | Experimental study | N/A | Glioma patients | Mass spectrometry | Onco-metabolite mapping | Real-time detection of 2-hydroxyglutarate for surgical guidance | Excellent |
| Artificial Intelligence | | | | | | | |
| Tampu et al. (2025) | Challenge study | 178 | Pediatric brain tumors | Deep Learning + MRI | Classification of pediatric brain tumors | 87% accurate with vision transformer model and age fusion | Excellent |
| Lee et al. (2024) | Federated learning study | 1247 | Pediatric brain tumors | Federated AI platform | Model generalization | Area under curve 0.85-0.92 across 19 international sites | Excellent |
| Alleman et al. (2023) | Systematic Review | N/A | Pediatric low-grade gliomas | Deep learning | Recurrence prediction | Superior risk stratification combining imaging and clinical variables | Excellent |
| Isensee et al. (2021) | Method development | 23 | Brain tumor patients | nnU-Net | Automated segmentation | Self-configuring network superior to manual segmentation methods | Excellent |
| Multi-modal Imaging | | | | | | | |
| Pan et al. (2023) | Classification study | 103 | Pediatric DIPG | Multi-modal MRI+PET | Tumor classification | Identified four distinct DIPG subtypes with different treatment implications | Good |
| Xiao et al. (2021) | DTI study | 54 | Brainstem lesions (pediatric) | DTI + conventional MRI | Surgical decision making | Modified surgical approach in 33.3% of cases based on tract visualization | Good |
