## Supplementary Table S1 for "Intraoperative Metabolomic-Guided Precision Surgery for Pediatric Brain Tumors: A Systematic Review of Multi-Modal Molecular Imaging Platforms and Artificial Intelligence Integration"

| Author (Year) | Design | Country | Sample Size | Population | Technology | Primary Outcome | Key Finding | Quality |
|---|---|---|---|---|---|---|---|---|
| Abernethy et al. (2012) | Retrospective cohort | UK | 55 | Pediatric brain tumors (2-17 years) | 3T iMRI | Extent of resection | GTR increased from 67% to 89% with iMRI | Good |
| Alleman et al. (2023) | Systematic review | USA | N/A | Pediatric low-grade gliomas | Deep learning | Recurrence prediction | Improved risk stratification | Excellent |
| Baid et al. (2021) | Benchmark study | USA | 2000 | Brain tumor patients | BraTS challenge | Segmentation benchmark | State-of-the-art performance | Excellent |
| Balog et al. (2013) | Technology development | UK | 91 | Various surgical patients | REIMS | Real-time identification | 2-3 minute analysis time | Good |
| Basu et al. (2021) | Clinical trial analysis | USA | 112 | Breast cancer patients | Multi-modal MS | Intraoperative guidance | Effective surgical guidance | Good |
| Bennett et al. (2018) | Metabolomic analysis | Spain | 83 | Pediatric cerebellar tumors | HR-MAS NMR | Metabolite profiling | Distinct tumor-specific signatures | Good |
| Borja et al. (2013) | Imaging study | Italy | 89 | Pediatric brain tumors | Advanced MRI | Diagnostic accuracy | MRI sequence optimization | Good |
| Bou-Samra et al. (2021) | Future perspective | USA | N/A | Molecular imaging | Intraoperative imaging | Technology roadmap | Promising future directions | Good |
| Calligaris et al. (2014) | Imaging study | USA | 85 | Breast cancer patients | DESI-MS imaging | Margin analysis | Real-time margin assessment | Excellent |
| Casey et al. (2005) | Development study | USA | 67 | Children/adolescents | Functional MRI | Cognitive development | Functional reorganization | Good |
| Cheng et al. (1997) | Spectroscopy study | USA | 45 | Brain pathology | HR-MAS NMR | Neuropathology quantification | Quantitative tissue analysis | Good |
| Choudhri et al. (2014) | Retrospective analysis | USA | 168 | Pediatric CNS neoplasms (0.5-18 years) | 3T iMRI | Residual tumor detection | Improved complete resection rates by 23% | Good |
| Clark et al. (2018) | Validation study | Canada | 8 | Medulloblastoma (pediatric) | Handheld MS | Subgroup classification | Rapid subgroup identification | Good |
| Claudino et al. (2012) | Review study | Brazil | 145 | Cancer patients | Cancer metabolomics | Metabolomic applications | Bench-to-bedside translation | Good |
| Coan et al. (2014) | Case series | France | 12 | Hippocampal lesions (pediatric) | iMRI | Diagnostic accuracy | Improved lesion characterization | Fair |
| Danilov et al. (2020) | Systematic review | USA | 143 | Neurosurgical AI applications | Various AI methods | Clinical applications | Rapid growth in AI adoption | Good |
| Duffau (2012) | Perspective study | France | 234 | Low-grade glioma | Functional mapping | Function preservation | Plasticity-based surgery | Excellent |
| Eberlin et al. (2013) | Prospective cohort | USA | 32 | Brain tumors (includes pediatric) | DESI-MS | Tissue classification accuracy | 96% accuracy for tumor detection | Excellent |
| Falco et al. (2021) | Technical study | UK | 78 | Pediatric patients | iMRI positioning | Technical feasibility | Reduced procedure complications | Good |
| Fiehn (2002) | Conceptual study | Germany | N/A | Metabolomics | Metabolomics methods | Genotype-phenotype link | Metabolomics framework | Good |
| Finlay et al. (1996) | Pilot study | USA | 34 | Recurrent astrocytoma (pediatric) | Image guidance | Treatment response | Improved targeting accuracy | Fair |
| Gajjar et al. (2014) | Case series | Austria | 28 | Glioblastoma patients | LITT + iMRI | Treatment efficacy | Precise thermal ablation | Good |
| Giussani et al. (2010) | DTI validation | Italy | 23 | Brainstem gliomas | DTI fiber tracking | Tract visualization | Improved surgical planning | Fair |
| Gogtay et al. (2004) | Longitudinal study | USA | 89 | Healthy children/adolescents | Structural MRI | Cortical development | Dynamic cortical maturation | Excellent |
| Haberg et al. (2016) | Population study | Norway | 1006 | General population | Incidental findings | Clinical significance | Incidental finding rates | Good |
| Hanihara et al. (2023) | Prospective cohort | UK | 24 | Posterior fossa tumors (1-16 years) | Low-field iMRI | Surgical decision modification | Additional resection in 34% of cases | Good |
| Higgins et al. (2011) | Methodology study | UK | N/A | RCT assessment | Risk of bias tool | Bias assessment | Systematic bias evaluation | Excellent |
| Isensee et al. (2021) | Method development | Germany | 23 | Brain tumor patients | nnU-Net | Automated segmentation | Superior to manual methods | Excellent |
| Jarmusch et al. (2016) | Analytical study | USA | 39 | Brain tumor patients | DESI-MS | Lipid profiling | Distinctive metabolic signatures | Excellent |
| Jehi et al. (2015) | Prediction study | USA | 435 | Epilepsy surgery patients | AI prediction models | Seizure outcome prediction | Individualized outcome prediction | Excellent |
| Jellema et al. (2023) | Systematic review | Netherlands | N/A | Pediatric brain tumors | Advanced iMRI | Clinical outcomes | Significant improvement in PFS with iMRI | Excellent |
| Joseph et al. (2025) | AI integration study | India | 89 | Neonatal surgery | AI algorithms | Surgical precision | Enhanced surgical outcomes | Good |
| Karsy et al. (2019) | Retrospective analysis | USA | 89 | Pediatric brain tumors | iMRI | Neurological outcomes | Preserved function in 87% | Good |
| Labuschagne (2020) | Retrospective | South Africa | 22 | Pediatric posterior fossa tumors | 5-ALA FGS | Extent of resection | 5-ALA assisted GTR in posterior fossa tumors | Good |
| LeCun et al. (2015) | Review article | USA | N/A | Deep learning | Neural networks | Technology overview | Deep learning foundations | Excellent |
| Lee et al. (2024) | Federated learning study | Multi-national | 1247 | Pediatric brain tumors | Federated AI platform | Model generalization | AUC 0.85-0.92 across sites | Excellent |
| Li et al. (2021) | AI validation study | USA | 156 | Pediatric brain tumors | Label-free molecular imaging | Recurrence prediction | Superior to conventional imaging | Excellent |
| Linguraru et al. (2010) | Segmentation study | USA | 156 | Abdominal imaging | Automated segmentation | Organ quantification | Reliable automated analysis | Good |
| Litjens et al. (2017) | Survey study | Netherlands | 300 | Medical imaging | Deep learning survey | Technology review | Comprehensive DL applications | Excellent |
| Merchant et al. (2010) | Multi-age analysis | USA | 789 | Brain tumors across ages | Age-stratified analysis | Age-specific outcomes | Age-dependent treatment | Excellent |
| Milos et al. (2023) | Prospective study | Sweden | 14 | Pediatric brain tumors (4-17 years) | 5-ALA + spectroscopic probe | Fluorescence detection | Vague microscopic fluorescence in 2/14; probe de | Good |
| Mobadersany et al. (2018) | Deep learning study | USA | 1053 | Glioma patients | CNN analysis | Survival prediction | Outperformed pathologists | Excellent |
| Mohammadi et al. (2014) | Multi-center study | USA | 67 | High-grade glioma | LITT therapy | Progression-free survival | Improved PFS with LITT | Good |
| Moher et al. (2009) | Guideline study | Canada | N/A | Systematic reviews | PRISMA methodology | Reporting guidelines | Improved review quality | Excellent |
| Ng et al. (2023) | Systematic review | USA | 89 | Pediatric medical imaging | AI applications | Diagnostic accuracy | Improved pediatric outcomes | Good |
| Ntenti et al. (2023) | Multi-center study | USA | 234 | Pediatric ependymoma | Molecular characterization | Prognostic factors | Molecular subtyping importance | Excellent |
| Ostrom et al. (2020) | Epidemiological study | USA | 12456 | CNS tumor registry | Population analysis | Incidence patterns | Updated epidemiology | Excellent |
| Öz et al. (2014) | Review study | USA | 287 | Neurological patients | MR spectroscopy | Diagnostic utility | Valuable diagnostic tool | Good |
| Pan et al. (2023) | Classification study | China | 103 | Pediatric DIPG | Multi-modal MRI+PET | Tumor classification | Four distinct DIPG subtypes | Good |
| Pekov et al. (2025) | Review article | Russia | 234 | Neurosurgical patients | Intraoperative MS | Clinical utility | Promising for pediatric applications | Good |
| Phelps et al. (2018) | Validation study | UK | 123 | Gynecological patients | REIMS | Tissue classification | Rapid tissue identification | Good |
| Pirro et al. (2017) | Clinical trial | USA | 81 | Glioma patients (adult/pediatric) | DESI-MS | Margin assessment | Superior to frozen section | Excellent |
| Pirro et al. (2017) | Application study | USA | 45 | Glioma patients | Touch spray MS | Margin assessment | Oncometabolite detection | Good |
| Pollack et al. (2019) | Comprehensive review | USA | 567 | Childhood brain tumors | Current management | Treatment outcomes | Integrated care benefits | Excellent |
| Raghib et al. (2022) | Retrospective cohort | USA | 89 | Repeat craniotomy cases (2-17 years) | iMRI guidance | Surgical outcomes | Reduced reoperation rates | Good |
| Rajpurkar et al. (2022) | Review article | USA | N/A | Healthcare AI | AI applications | Healthcare AI review | Broad healthcare applications | Excellent |
| Ren et al. (2020) | Connectome study | China | 67 | Brain tumor patients | Structural/functional MRI | Connectome analysis | Personalized therapy guidance | Good |
| Ronneberger et al. (2015) | Method development | Germany | 347 | Medical imaging | U-Net architecture | Segmentation accuracy | Superior biomedical segmentation | Excellent |
| Santagata et al. (2014) | Experimental study | USA | N/A | Glioma patients | Mass spectrometry | Onco-metabolite mapping | Real-time 2-HG detection | Excellent |
| Sawamura et al. (2008) | Case series | Germany | 34 | Hypothalamic astrocytomas (3-16 years) | iMRI | Hypothalamic preservation | 91% functional preservation | Fair |
| Schwake et al. (2019) | Systematic review | Germany | N/A | Pediatric brain tumors | 5-ALA FGS | Fluorescence utility | Helpful in 78% GBM, 71% anaplastic ependymom | Excellent |
| Shen et al. (2017) | Review article | USA | N/A | Medical imaging | Deep learning | Method analysis | Promising medical applications | Excellent |
| Sievert & Fisher (2009) | Review study | USA | 156 | Pediatric LGG | Multi-modal assessment | Management strategies | Comprehensive care approach | Good |
| Stummer et al. (2006) | Randomized controlled tria | Germany | 322 | Malignant glioma (adult) | 5-ALA FGS | Complete resection rate | 65% GTR with 5-ALA vs 36% with white light; lan | Excellent |
| Sun et al. (2021) | Systematic Review | USA | N/A | Glioma patients | 5-ALA FGS | Fluorescene utility and rationale | Improved extent of resection and prognosis | Good |
| Tampu et al. (2025) | Challenge study | USA | 178 | Pediatric brain tumors | CNN segmentation | Segmentation accuracy | Dice coefficient 0.78-0.89 | Excellent |
| Tejada et al. (2018) | Cohort study | Spain | 156 | Glioblastoma (adult/pediatric) | iMRI | Complete resection rate | Higher GTR with iMRI guidance | Good |
| Thompson et al. (2019) | Retrospective analysis | UK | 56 | Craniopharyngioma (2-17 years) | iMRI | Hypothalamic morbidity | Reduced endocrine dysfunction | Good |
| Topol (2019) | Perspective article | USA | N/A | Healthcare | AI convergence | Future medicine | AI-human convergence | Excellent |
| Traylor et al. (2021) | Prospective Study | USA | 23 | GBM patients | Laser therapy + iMRI | Technical feasibility | Precise ablation guidance | Good |
| Traylor et al. (2021) | Review article | USA | N/A | Glioma patients | 5-ALA metabolic mechanism | PpIX fluorescence mechanism | Characterized molecular and metabolic mechanis | Excellent |
| Vaysse et al. (2020) | Clinical study | UK | 95 | Breast surgery patients | REIMS | Pathology identification | Intelligent knife capability | Good |
| Wang et al. (2024) | Systematic review | China | N/A | Pediatric brain tumors (27 studies) | 5-ALA FGS | Fluorescence rate and safety | Fluorescence rate >75% in high-grade tumors; sa | Excellent |
| Wataya (2017) | Conference abstract | Japan | N/A | Pediatric brain tumors | 5-ALA FGS | Fluorescence utility | Age-dependent fluorescence patterns in pediatric | Fair |
| Wells et al. (2000) | Assessment tool | Canada | N/A | Observational studies | Quality assessment | Study quality evaluation | Standardized quality tool | Good |
| Wilson et al. (2009) | Prospective analysis | UK | 57 | Childhood brain tumors | 1H NMR spectroscopy | Metabolic characterization | Age-specific metabolic patterns | Good |
| Wilson et al. (2019) | Consensus study | Multi-national | 456 | Brain MRS patients | Proton MRS | Methodological consensus | Standardized protocols | Excellent |
| Woolman et al. (2017) | Method validation | Canada | 67 | Cancer patients | MS workflow | Metabolite identification | Optimized analysis protocol | Good |
| Wu et al. (2024) | Literature review | Australia | 423 | Pediatric brain tumors | iMRI | Resection outcomes | Consistent improvement across studies | Good |
| Wu et al. (2024) | Literature analysis | Australia | 234 | Pediatric iMRI | iMRI evaluation | Clinical utility | Consistent benefit across studies | Good |
| Xiao et al. (2022) | DTI study | UK | 54 | Brainstem lesions (pediatric) | DTI + conventional MRI | Surgical decision making | Modified approach in 73% cases | Good |
| Xue et al. (2018) | Retrospective | China | 12 | Pediatric brainstem gliomas (2-18 years) | Sodium fluorescein | Resection extent and safety | 100% fluorescence utility; GTR in 75%; safe dose | Good |
| Yang et al. (2025) | AI application study | China | 234 | Nasopharyngeal carcinoma | AI radiotherapy | Treatment planning | Improved treatment accuracy | Good |
| Zhou et al. (2021) | Review study | USA | 789 | Medical imaging | Deep learning | Technology trends | Rapid advancement in medical AI | Excellent |
